# Negative attitudes about facemasks during the COVID-19 pandemic: The dual importance of perceived ineffectiveness and psychological reactance

**DOI:** 10.1101/2020.11.17.20233585

**Authors:** Steven Taylor, Gordon J. G. Asmundson

## Abstract

This study reports a comprehensive empirical investigation of the nature and correlates of anti-mask attitudes during the COVID-19 pandemic. Accumulating evidence underscores the importance of facemasks, as worn by the general public, in limiting the spread of infection. Accordingly, mask-wearing has become increasingly mandatory in public places such as stores and on public transit. Although the public has been generally adherent to mask-wearing, a small but vocal group of individuals refuse to wear masks. Anti-mask protest rallies have occurred in many places throughout the world, sometimes erupting violently. Few empirical studies have examined the relationship between anti-mask attitudes and mask non-adherence and little is known about how such attitudes relate to one another or other factors (e.g., non-adherence to social distancing, anti-vaccination attitudes). To investigate these issues, the present study surveyed 2,078 adults from the US and Canada. Consistent with other surveys, we found that most (84%) people wore masks because of COVID-19. The 16% who did not wear masks scored higher on most measures of negative attitudes towards masks. Network analyses indicated that negative attitudes about masks formed an intercorrelated network, with the central nodes in the network being (a) beliefs that masks are ineffective in preventing COVID-19, and (b) psychological reactance (PR; i.e., an aversion to being forced to wear masks). These central nodes served as links, connecting the network of anti-masks attitudes to negative attitudes toward SARSCoV2 vaccination, beliefs that the threat of COVID-19 has been exaggerated, disregard for social distancing, and political conservatism. Findings regarding PR are important because, theoretically, PR is likely to strengthen other anti-masks attitudes (e.g., beliefs that masks are ineffective) because people with strong PR react with anger and counter-arguments when their beliefs are challenged, thereby leading to a strengthening of their anti-mask beliefs. Implications for improving mask adherence are discussed.

## Introduction

### Adherence and Non-Adherence to Wearing Facemasks

Historically, protective facemasks have played an important role in limiting the spread of infection during pandemics (1). Mask-wearing has long been regarded as vital for healthcare workers and has been recommended for people in the community, although there have been inconsistent messages from health authorities about mask-wearing for the general public during the COVID-19 pandemic (2-4). Early in the pandemic, some health authorities such as the World Health Organization (WHO) recommended that masks not be worn by the public and that masks should be reserved for health care workers and those infected with COVID-19 (4, 5). Early in the pandemic the WHO and other health authorities also averred that masks are ineffective for the general public, that masks provided a false sense of security, and that the wearing of masks would cause people to touch their faces, thereby increasing their chances of infection (4, 6). Such concerns proved to be unfounded (6-8) and evidence steadily emerged to support the wearing of masks by members of the general public (9, 10). Health authorities such as the WHO subsequently reversed their position on facemasks and recommended that masks be worn by the public when they were in public places, especially when social distancing was not possible (5).

The prevalence of mask-wearing in the community increased as COVID-19 spread globally (5, 11). In the US and Canada, which were the counties in which the present research was focused, mask-wearing in the community increased dramatically in the months following the formal declaration on March 11, 2020 that COVID-19 had become a pandemic (12-15). Surveys conducted from June to August, 2020 suggested that most (approximately 80-85%) of Canadian and American adults regularly wore masks (5, 13-16). The increase in mask-wearing was likely due to several factors including (a) the increasing prevalence of COVID-19, (b) reversal by health authorities on their earlier position, from dissuading to encouraging the public to wear masks, and (c) mask-wearing had become increasingly mandatory in stores and on public transport. In the months after the pandemic was declared, the policy of mandatory masks appeared to gain acceptance among the general public. Surveys in July, 2020 suggested that most respondents (74%) found that a policy of mandatory mask-wearing was personally acceptable (13), which was an increase from 37% in May (12).

Despite the importance of masks, some people object to wearing them. During the so-called Spanish flu pandemic (1918-1920), for example, opposition to wearing masks occurred in San Francisco in 1919, when authorities attempted to make it mandatory to wear masks in public places. The Anti-Mask League was formed, which was a short-lived protest movement in which the proponents argued that masks were ineffective, inconvenient, and that mandatory mask-wearing violated their civil liberties (17, 18). These anti-maskers were a vocal but small group, so most people followed the recommendations to wear masks in public (17, 18).

A similar situation has arisen during the COVID-19 pandemic. Estimates suggest that at the time the present research was conducted (July-August, 2020), about 10-15% of adults in the US and Canada rarely or never wore masks in public (13, 15, 16). During the COVID-19 pandemic, there have been numerous anti-mask protest rallies, primarily in the US, Canada, Europe, and Australia, with some rallies drawing thousands of people (19-22). Protest rallies have persisted despite high rates of infection and large numbers of deaths from COVID-19. Violence has erupted in some rallies as protesters clashed with counter-protesters or with police (20, 22-25). Just as in 1919, the COVID-19 anti-mask protests were spurred by government plans to make masks mandatory. Some rallies were protests against mandatory masks, while others were broader, protesting against masks, social distancing, lockdown, and vaccines (21, 22, 26). Protesters at some anti-masks rallies were joined by people espousing conspiracy theories, such as the unfounded belief that the threat of COVID-19 has been exaggerated by the government in order to control the populace (27).

Unlike the 1919 anti-mask movement, which was short-lived and limited to a single city, the anti-mask rallies during COVID-19 have taken place in many cities, sporadically arising without any evidence (at the time of writing) of petering out. The recurrence of such rallies is likely fueled by social media and news media coverage combined with strongly engrained anti-mask attitudes held by at least a small proportion of the population.

Very little is known about the motivations for mask non-adherence. Public opinion polls suggest that people with conservative political affiliations (Republicans in the US or Conservatives in Canada) are less likely to wear masks than people with liberal affiliations (Democrats or Liberals) (5, 14, 16, 28). This may be partly because Republican political leaders were initially reluctant to wear masks, even mocking those who wore masks (5, 16), and because people with politically conservative ideologies tend to resist government regulatory efforts (29), such as attempts to make masks mandatory.

Several opinion polls have assessed reasons for not wearing masks, although the polls were limited in the number of reasons assessed. The most common of the assessed reasons for not wearing masks were: Not believing that masks are effective, finding masks uncomfortable, difficulty establishing the habit of mask-wearing, and lack of concern about COVID-19 (13, 14, 16, 30). Newspaper reports of protest rallies offer additional, anecdotal information on anti-masks sentiments. Several reasons for not wearing masks were suggested by news reports of anti-mask protests: Beliefs that masks violate civil liberties, beliefs that masks are ineffective and possibly harmful because masks make breathing difficult, and beliefs that the threat of COVID-19 has been exaggerated (2, 19, 21, 22, 31-37). Despite the range of anti-mask attitudes, a common theme running through these reports is that protestors believe that mandatory masks are a violation of civil rights. In other words, anecdotal news reports suggest that the rallies are motivated, at least in part, by a phenomenon known as psychological reactance (PR).

### Psychological Reactance

People like to feel in control (38). Relatedly, PR is a motivational response to rules, regulations, or attempts at persuasion that are perceived as threatening one’s sense of control, autonomy, or freedom of choice (39, 40). The perceived threat motivates the person to assert their freedom by rejecting attempts at persuasion, rules, regulation, and other means of control. Thus, when PR is evoked it is characterized by counter-arguments and anger (41). This might involve denying the existence of a threat (39, 42); for example, denying the need to wear masks by denying the seriousness of the pandemic. Thus, PR is expected to be correlated with a denial or disregard for the seriousness of the COVID-19 pandemic.

The tendency to experience PR is a personality trait (43), which is correlated with a range of phenomena including antisocial and narcissistic personality traits (44, 45) and with political conservatism (46, 47). The latter finding is not surprising given that conservative ideology, as compared to liberal ideology, strongly favors limited government intervention (29), and so government policies that threaten to restrict freedoms are likely to elicit PR (46). Mass communication messages that try to persuade people to adopt a given behavior, such as wearing masks, can elicit PR, thereby undermining the impact of the message (40, 48). Given these considerations, it is important to investigate whether PR is implicated in mask non-adherence during the COVID-19 pandemic.

### Aims of the Present Study

Although most people have been adherent to wearing masks during the COVID-19 pandemic, opinion polls suggest that a small but significant proportion of people (10-15%) object to wearing masks. News reports, although anecdotal, suggest that people who object to wearing masks are a vocal minority, engaging in protest rallies that sometimes erupt in violence. Accordingly, non-adherence to wearing masks is a socially important phenomenon as well as one that is relevant to managing the COVID-19 pandemic. To date, the study of mask non-adherence during the COVID-19 pandemic has been limited to a small number of opinion polls, with a limited assessment of anti-masks attitudes. The present study had three primary aims.

The first aim was to conduct a broader examination of anti-mask attitudes, to determine which attitudes are related to masks non-adherence, and to investigate how anti-mask attitudes are related to one another. Previous research on PR suggests that mask-related PR (i.e., objecting to being forced to wear masks) should be related to other anti-mask attitudes, because when a person with a high degree of PR is challenged regarding a given attitude (e.g., the belief that masks are ineffective), then that person will resist the efforts at belief change by generating counter-arguments to support the attitudes (e.g., generating counter-arguments to support the belief that masks are ineffective).

The second aim was to investigate how anti-mask attitudes are related to political conservatism. Previous research, as discussed above, suggests that the general propensity to PR is related to conservatism; but, it has yet to be empirically established whether mask-related PR is related to political conservatism. The final aim was to investigate how anti-mask attitudes are related to attitudes and behaviors that are associated with non-adherence to other pandemic-control measures during the COVID-19 pandemic, such as disregard for social distancing and anti-vaccination attitudes. Recent research provides evidence for a COVID-19 disregard “syndrome” (49). This is not a syndrome in the medical sense of the term, but rather a constellation of inter-related attitudes and beliefs. People with this syndrome tend to (a) believe that the COVID-19 pandemic has been exaggerated, (b) see themselves as physically robust to any illness they may experience as a result of being infected with SARSCoV2, and (c) tend to disregard social distancing because they see it as unnecessary (49). This syndrome is also associated with negative attitudes toward a potential vaccine for SARSCoV2 (i.e., beliefs that a vaccine is unnecessary or that the benefits of such a vaccine are outweighed by the potential risks) (49).

The first aim of this study was addressed by simple t-tests comparing anti-mask attitudes of people who were adherent versus non-adherent to wearing masks. The remaining aims were address by network analyses, which are well-suited for gaining insights into the complex interplay among variables. Network analysis provides important information about relationships among elements (nodes) in a network (e.g., sets of attitudes or behaviors). Network analysis assumes that nodes are inter-related because they are, in some way, causally linked with one another. In network analysis, the links are known as “edges.” The presence of statistically significant edges does not assume that nodes are influenced by some underlying factor such as a latent variable. Instead, network analysis assumes that nodes may directly influence one another (50). In the present study, anti-mask attitudes were predicted to form a network of interconnected nodes. Given that PR theoretically amplifies the strength of other attitudes, mask-related PR was predicted to be one of the central nodes in the network of mask-related attitudes. Network analyses were also conducted to determine how the network of anti-mask attitudes is related to other variables: Political conservativism, the elements of the COVID-19 disregard syndrome, and SARSCoV2 anti-vaccination attitudes. If nodes causally influence one another, then changes in a central node are most likely to lead to changes in other nodes in the network through the spreading of activation. Central nodes, as compared to peripheral nodes, are defining features of a network. Identifying central nodes has the potential to inform which elements to target in interventions. As a caveat, note that network analyses in cross-sectional designs such as the present study are suggestive of, but do not establish causality. Significant edges might represent causal influences (either unidirectional or directional) but experimental designs are needed to establish causality. Accordingly, network analyses provide a source of hypotheses about causal links among variables in a network.

## Method

### Sample

The sample consisted of 2,078 adults (age ≥18 years) from the United States (*N*=1,036) and Canada (*N*=1,042). The mean age was 54 years (SD=14 years, range 18-94 years). Most (93%) were employed full- or part-time, most (82%) had completed full or partial college, and 40% were female. Most (70%) were Caucasian, with the remainder being Asian (13%), African American/Black (8%), Latino/Hispanic (4%), or other (4%). Only 2% of the sample reported being diagnosed with COVID-19.

### Data Collection Procedures

Data were collected from July 20 to August 7, 2020, using an internet-based self-report survey delivered in English by Qualtrics, a commercial survey sampling and administration company. Qualtrics solicited the present sample as part of our ongoing research program (51, 52). Qualtrics maintains a pool of potential participants who have agreed to be contacted in order to respond to surveys. Qualtrics selected and contacted participants to meet sampling quotas based on age, gender, ethnicity, socioeconomic status, and geographic region within each country. All respondents provided written informed consent prior to completing the survey. The research described in this article was approved by the Research Ethics Board of the University of Regina (REB# 2020-043). Filters were used to eliminate data from careless responders. Embedded in the assessment battery were four attention-check items (e.g., “This is an attention check, please select Strongly Agree”; “For our research, it is really important that you paid attention while responding to our survey. How attentive were you when responding?”: “Very Inattentive” to “Very Attentive”). Participants were included only if they provided correct responses to three or more of the four attention checks (e.g., “Strongly agree” or “Very attentive”), indicating that they were sufficiently attentive. In addition, at the end of the assessment battery, participants were asked to indicate whether, in their honest opinion, their data should be used. Those who responded “no” were excluded from data analysis, regardless of their score on the attention-check items.

### Measures

Participants completed a battery of measures, including demographic questions. Adherence to wearing facemasks was assessed with a face-valid yes/no item: “Do you wear a facemask because of concerns about COVID-19?” Participants completed a 12-item scale, developed for the purpose of the present study, assessing negative attitudes about facemasks. The items, derived from previous descriptions of anti-mask attitudes (1, 13, 14, 16-18, 30, 53), are listed in Table 1. Each item was rated on a 7-point scale (1=strongly disagree, 7=strongly agree). Three scales, previously developed to assess the COVID Disregard Syndrome (49), were also administered: (a) Belief that the dangerousness of COVID-19 is exaggerated, (b) disregard for social distancing, and (c) belief that one has robust personal health against infection. Items on these scales were rated on a 5-point scale (0=strongly disagree, 4=strongly agree). These face-valid scales have good levels of reliability and validity (49). Anti-vaccination attitudes toward a SARSCoV2 vaccine were measured using an adaptation of the Vaccination Attitudes Examination Scale (54), assessing vaccination attitudes specific to SARSCoV2 (55). The items in this scale, each rated on a 6-point scale (0=strongly disagree, 5=strongly agree), assess mistrust of vaccine benefit, worries over unforeseen future effects of the vaccine, concerns about commercial profiteering from the vaccine, and preference for natural immunity. The scale has good levels of reliability and validity (54, 55). Political conservatism was assessed with a single face-valid item: “In general, how would you describe your political views?” (1=very liberal, 7=very conservative).

**Table 1.** Scores on measures of negative attitudes towards facemasks.

|  | Figure 1<br>cluster | People<br>who<br>don't<br>wear<br>masks<br>(N=338)<br>M (SD) | People<br>who<br>wear<br>masks<br>(N=1,740)<br>M (SD) | t(df=2,026) |
| --- | --- | --- | --- | --- |
| I do not like feeling forced to wear a facemask (psychological reactance). | -- | 3.7 (2.3) | 3.0 (2.1) | 5.87*** |
| Facemasks are ineffective. | A | 3.3 (2.0) | 2.6 (1.7) | 6.53*** |
| Facemasks provide a false sense of security. | A | 4.0 (2.0) | 3.4 (1.9) | 5.66*** |
| Facemasks are unsafe because they force you to touch your face. | A | 3.2 (1.9) | 2.7 (1.7) | 5.22*** |
| It is hard to develop the habit of wearing a facemask. | B | 3.8 (2.1) | 3.0 (1.9) | 6.43*** |
| Wearing a facemask is too much of a hassle. | B | 3.2 (2.1) | 2.7 (1.8) | 5.15*** |
| Facemasks look ugly or weird. | C | 3.2 (2.0) | 2.9 (1.9) | 3.10** |
| Facemasks look silly. | C | 3.3 (2.1) | 3.0 (1.9) | 3.03** |
| Facemasks make other people feel uneasy. | D | 2.9 (1.9) | 2.6 (1.8) | 3.42*** |
| Facemasks make people look untrustworthy. | D | 3.2 (1.9) | 2.9 (1.8) | 2.54* |
| It is difficult to breathe when wearing a facemask. | E | 4.5 (1.9) | 4.3 (1.9) | 1.70 |
| Facemasks cause me to overheat. | E | 4.3 (1.9) | 4.2 (1.9) | 1.39 |

\* $p < .01$ , \*\* $p < .005$ , \*\*\* $p < .001$ .

### Statistical Procedures

For the network analyses, Glasso networks (regularized partial correlation networks) were computed using the R *qgraph* package (56). Indices of centrality, also calculated with *qgraph*, were used to assess the relative importance of each node in the network (57). Three indices of centrality were computed: *Strength, betweenness, and closeness. Strength* refers to how well a node is directly connected to other nodes in the network. Node strength is computed as the sum of the absolute values of edge weights (regularized partial correlations) that directly connect that node with other nodes. *Closeness* refers to how well a node is indirectly connected to other nodes in the network. Closeness is calculated by computing, for a given node, the inverse sum of edge weights for the shortest path between that node and each other node, and then summing the values for these paths. *Betweenness* refers to how important a given node is in the average path between two other nodes; that is, how often a given node is the most efficient (shortest) path between other nodes. This is an index of the importance of a given node in connecting nodes with one another. The stability (reliability) of the relative order of magnitude of edge weights and their strengths were tested by the correlation of stability coefficient, also calculated via *bootnet* (50). Coefficients exceeding .50 suggest stable (reliable) results (50). Due to the number of statistical tests conducted in this study, the α level was set at .01 instead of the conventional .05.

## Results

The majority (84%) of respondents reported that they wore facemask because of personal concerns about COVID-19, which indicates a high degree of adherence to public health recommendations. Negative attitudes about masks were largely uncorrelated with demographic variables (see S1 Appendix). Wearing of masks because of concerns about COVID-19 was also largely unrelated to demographic variables (see S1 Appendix). That is, mask-wearing (coded 1=yes, 0=no) was uncorrelated with age, gender, education level, and employment status (*r*s ranged from -.03 to .02; see S1 Appendix). Mask-wearing was significantly correlated with ethnic minority (non-White) status; but, the correlation was very small (*r*=.07, *p*<.001). Mask-wearing also had a small but significant correlation with country (coded as 1=Canada, 2=US); *r*=.13, *p*<.001). That is, significantly more people from the US than Canada reported wearing masks (90% vs. 78%). This may reflect the significantly higher per capita prevalence of COVID-19 disease and mortality in the US than Canada at the time the study was conducted (i.e., late July/early August). At that time, Canada was classified as a moderate mortality country and the US a high mortality country, with the estimated number of deaths per 100,000 people being 25 for Canada and 60 for the US (58).

Table 1 provides details about the degree of endorsement of negative attitudes about masks. People who did not wear masks because of COVID-19 tended to have more negative attitudes about masks as compared to people who wore masks. People who did not wear masks were most likely to report that they did not like being forced to wear a facemask (i.e., mask-related PR), believed that masks were ineffective and possibly harmful, believed that masks had adverse interpersonal effects, found masks to be esthetically unappealing, and found mask-wearing to be an inconvenient habit to form (see Table 1).

Anti-mask attitudes were strongly correlated with one another, with *r*s ranging from .35 to .91 and a mean *r* of .55 (*p*s<.001; see S1 Appendix). Figures 1 to 3 summarize the results of the network analyses. Figure 1 depicts the edges (regularized partial correlations) between nodes in the network of anti-mask attitudes. Edge values and their significance levels are shown in S1 Appendix. The figure shows five sets of strongly clustered attitudinal variables, as indicated by the strongest edges (i.e., thickest of the green connecting lines) in Figure 1: (A) Beliefs that masks are ineffective and possibly harmful, (B) beliefs that mask-wearing is an inconvenient habit to form, (C) beliefs that masks are esthetically unappealing, (D) beliefs that masks have adverse interpersonal effects, and (E) beliefs about the physical inconvenience of masks (i.e., difficulty breathing and overheating). As shown in Table 1, clusters A-D discriminated people who wore masks from those who did not. Figure 1 shows that at the center of the network was mask-related PR, which was connected to all clusters except cluster E. The coefficients of stability for the network shown in Figure 1 were .96 for edge weights and .75 for node strengths. These values exceed the cutoff of .50, suggesting stable (reliable) results.

**Figure 1.**
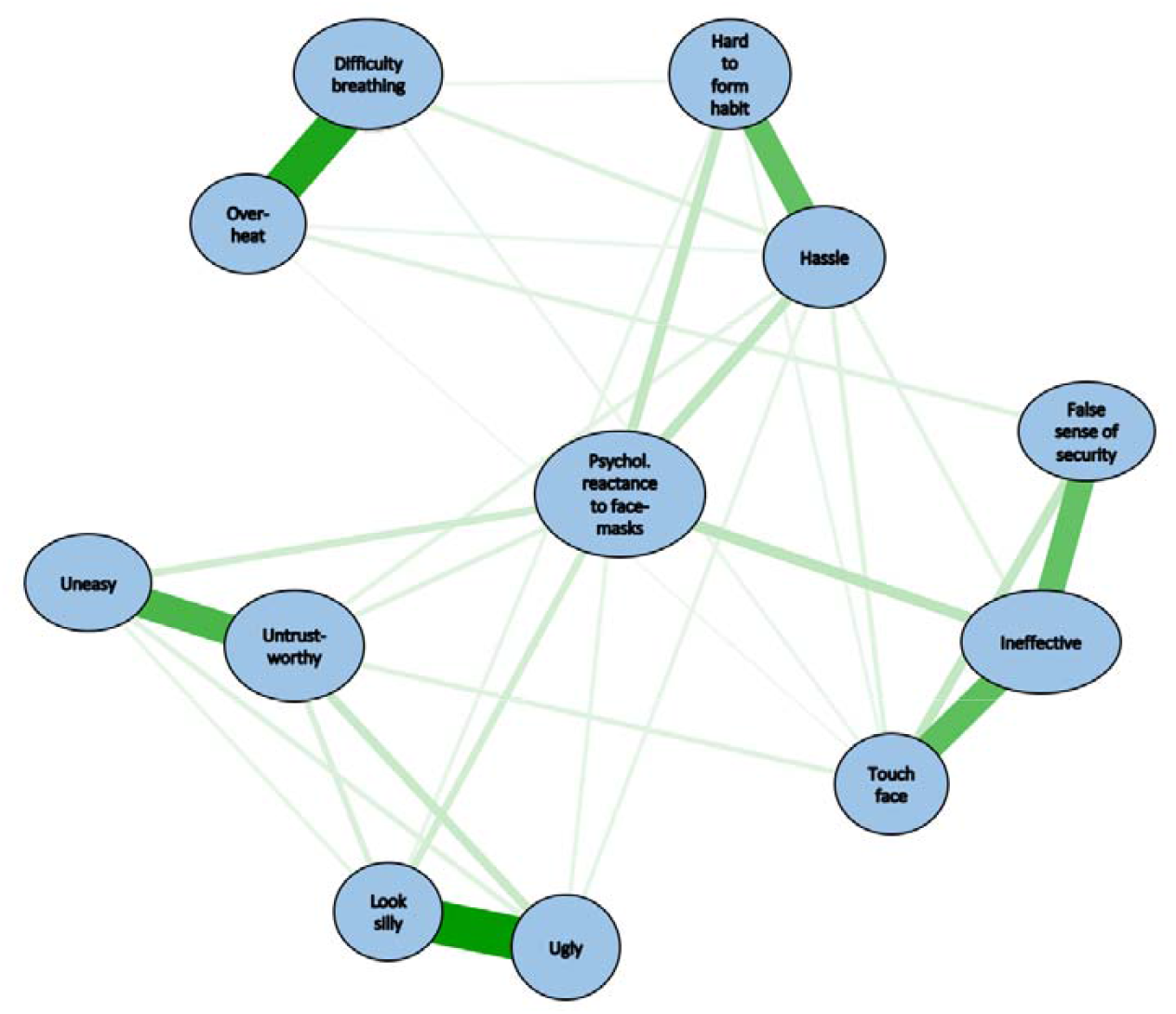
Network analysis of anti-mask attitudes: Strength of interconnections (edges) among the nodes in the network. All edges were positive (i.e., positive regularized partial correlations). Stronger edges are indicated by thicker lines. For all edges, *p*<.01. *Legend: Difficulty breathing*: It is difficult to breathe when wearing a facemask. *False sense of security*: Facemasks provide a false sense of security. *Hard to form habit:* It is hard to develop the habit of wearing a facemask. *Hassle:* Wearing a facemask is too much of a hassle. *Ineffective*: Facemasks are ineffective. *Look silly:* Facemasks look silly. *Overheat*: Facemasks cause me to overheat. *Psychol. reactance to facemasks:* I do not like feeling forced to wear a facemask. *Touch face:* Facemasks are unsafe because they force you to touch your face. *Ugly:* Facemasks look ugly or weird. *Uneasy:* Facemasks make other people feel uneasy. *Untrustworthy:* Facemasks make people look untrustworthy.

**Figure 2.**
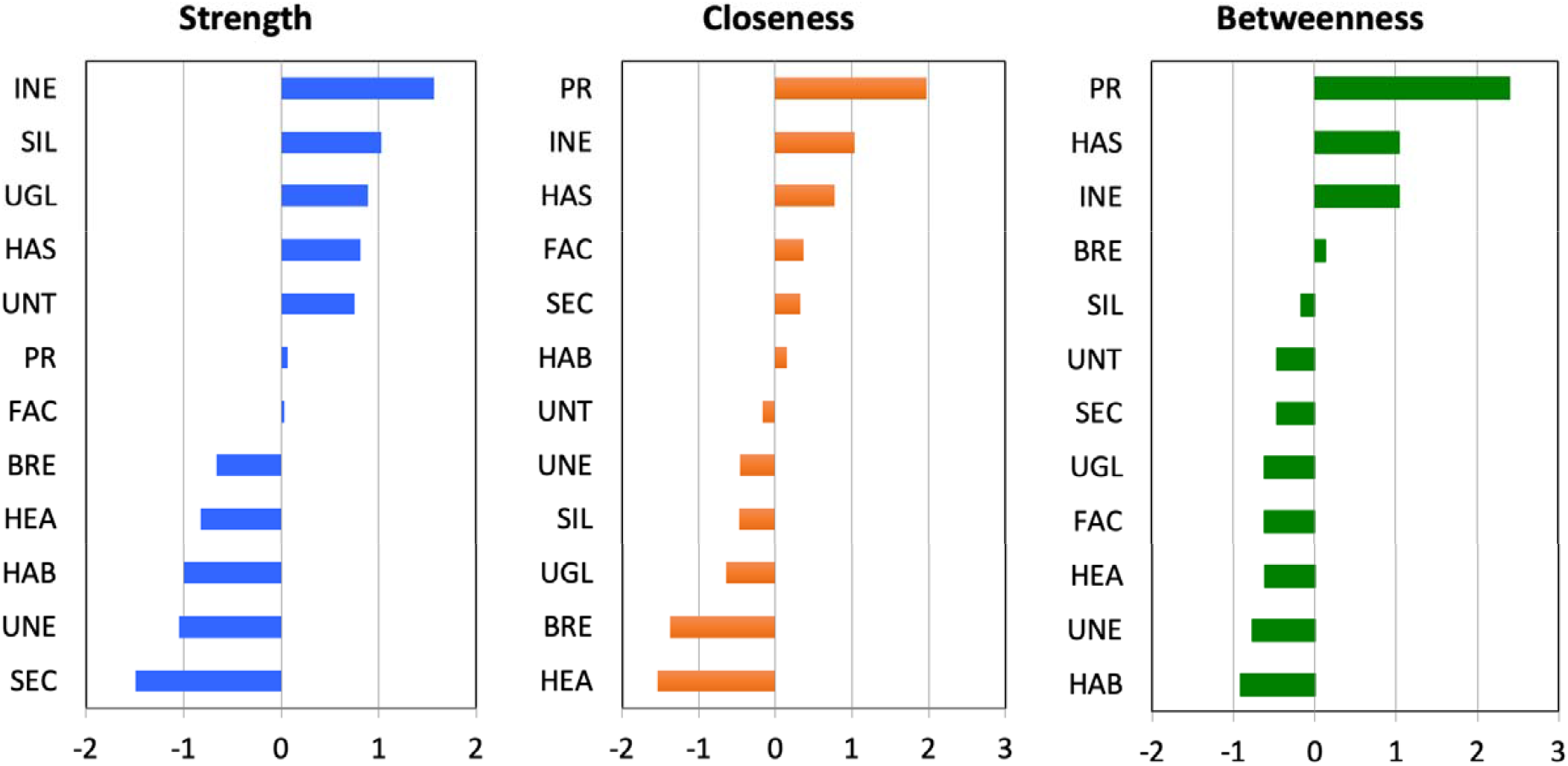
Centrality indices for nodes in the network analysis of anti-mask attitudes. Large values indicate that a given node had greater importance in the network, as indicated by its connections (edges) with other elements in the network. *Legend: BRE:* It is difficult to breathe when wearing a facemask. *FAC:* Facemasks are unsafe because they force you to touch your face. *HAB:* It is hard to develop the habit of wearing a facemask. *HAS:* Wearing a facemask is too much of a hassle. *HEA*: Facemasks cause me to overheat. *INE:* Facemasks are ineffective. *PR:* I do not like feeling forced to wear a facemask. *SEC:* Facemasks provide a false sense of security. *SIL:* Facemasks look silly. *UGL:* Facemasks look ugly or weird. *UNE:* Facemasks make other people feel uneasy. *UNT:* Facemasks make people look untrustworthy.

**Figure 3.**
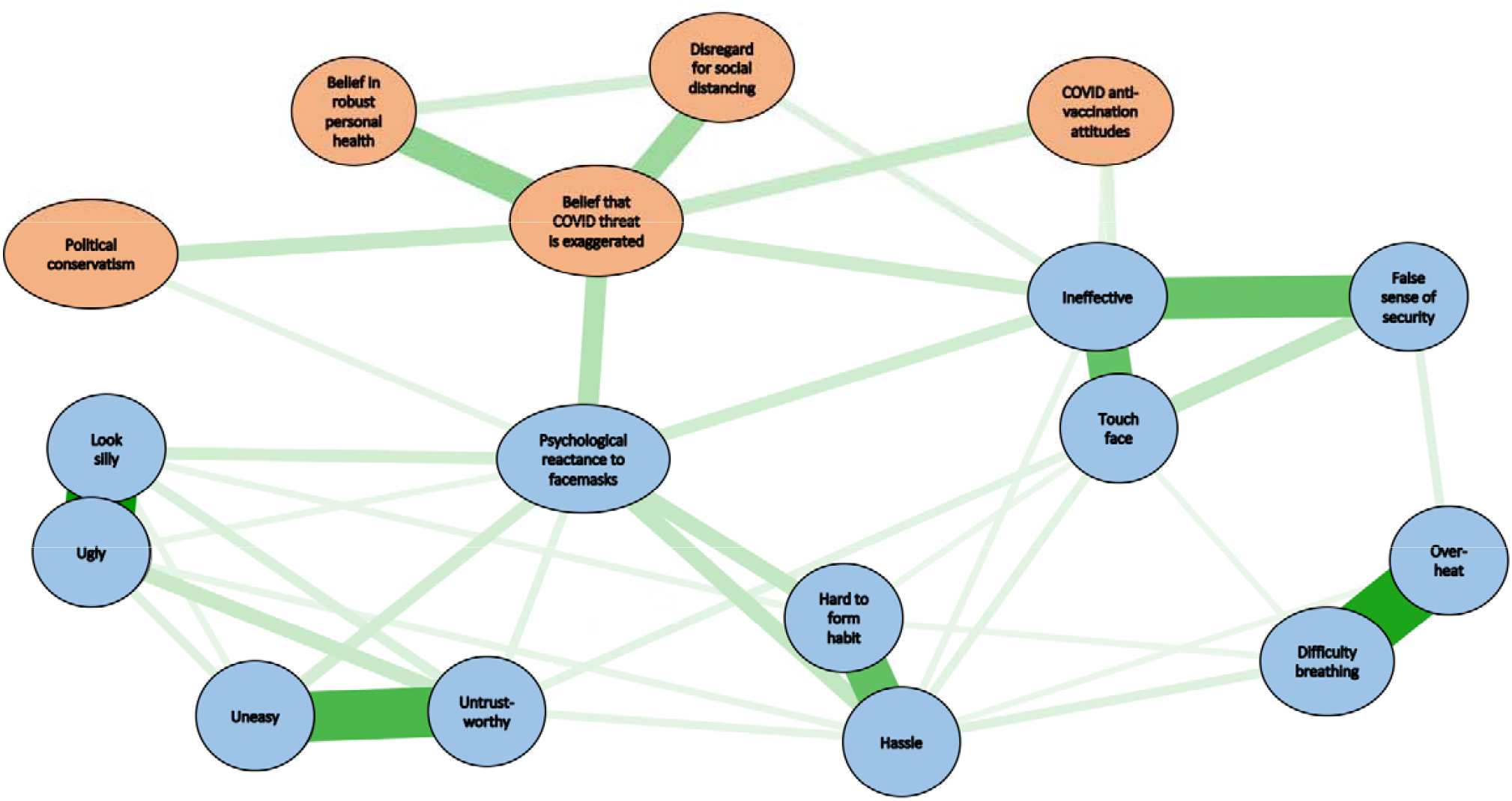
Extended network analysis linking anti-mask attitudes, COVID-19-disregard, and other variables. All edges were positive. Stronger edges are indicated by thicker lines. For all edges, *p*<.01.

Figure 2 shows the indices of centrality for the network in Figure 1. Figure 2 shows that beliefs that masks are ineffective was the strongest node in the network in that it was the node that was most directly connected to other nodes in the network. Figure 2 further shows that PR had the highest values on the closeness and betweenness indices. In other words, PR was highly important in terms of connecting other nodes with one another. In summary, Figures 1 and 2 indicate that PR and beliefs that masks are ineffective were the most important nodes in the network of anti-mask attitudes.

Figure 3 shows how the network of anti-mask attitudes was linked to other variables. For this network the coefficient of stability was .93 for edge weights, suggesting that the estimation of these values was stable (reliable). The actual values of the edge weights and their significance levels appear in S1 Appendix. For the network in Figure 3, centrality indices were not computed because they were not relevant to the aims of this analysis, which was to determine how the network of anti-mask attitudes was linked to other variables. The figure shows that the network of anti-mask attitudes was linked to the other variables by means of the two most important nodes in the anti-mask network—PR and beliefs that masks are ineffective. Beliefs that masks are ineffective was also linked to the nodes comprising the COVID Disregard Syndrome, and to COVID-19 anti-vaccination attitudes. PR was linked to the COVID Disregard Syndrome and to political conservatism. PR was significantly correlated with political conservatism for samples from both the US (*r*=.36, *p*<.001) and Canada (*r*=.28, *p*<.001), with no significant difference between the two at the α level of .01 (*z*=1.98, *p*>.04).

## Discussion

People who object to wearing masks are a small but highly vocal minority of individuals. The media attention that they have drawn may have given the misleading impression that the anti-mask sentiment is widespread. However, our findings, just like the findings from opinion surveys (5, 13-16) show that the majority of people are willing to follow the advice of health authorities about wearing masks. Refusal to wear masks is associated with a range of anti-mask attitudes, at the center of which are PR and beliefs that masks are ineffective. The core reasons identified in the present study are remarkably similar to the reasons voiced in 1919 by the Anti-Mask League; that is, they believed that masks were ineffective and violated their civil liberties. Figure 2 shows that the network of anti-mask attitudes is linked, via its central nodes (mask-related PR and beliefs that masks are ineffective), to other variables such disregard for social distancing and anti-vaccination attitudes. These findings underscore the importance of mask-related PR and beliefs that masks are ineffective.

A critical question for psychologists and those involved in public health messaging is, “What is the best way of encouraging people with anti-mask attitudes to wear masks?” It is important to consider the motivational roots of mask refusal. If a person refuses to wear a mask simply because he or she believes them to be ineffective, then it might be that targeted education may be sufficient. However, given that the belief that masks are ineffective is strongly correlated with mask-related PR (*r*=.66, *p*<.001; see S1 Appendix), this approach may be insufficient in many or perhaps most other cases. People who believe masks are ineffective also tend to have high levels of PR. If one tries to persuade a person with high PR that masks are effective, this will elicit reactance in which that person generates further arguments against the effectiveness of masks. The same applies to other reasons for mask refusal (e.g., the belief that masks make people look suspicious). Attempts to counter the majority of reasons for mask refusal will also elicit reactance because mask-related PR lies at the heart of the network of anti-mask attitudes.

PR is strongly correlated (*r*=.51) with psychopathic personality traits such as a tendency toward angry, impulsive behavior (45). Accordingly, confronting people who refuse to wear masks could be hazardous. Indeed, there have been media reports of people reacting violently when confronted about not wearing masks. For example, on public transit or in stores in which mask-wearing is mandatory, people who refuse to wear masks have verbally and physically assaulted people who have asked them to don a mask (59, 60). Accordingly, the CDC has recommended against confronting non-mask-wearers (61).

Alternative strategies are required, particularly persuasion strategies that address PR. Targeting PR is important because reactance lies at the heart of anti-mask attitudes. If the links in Figure 1 have any causal status, then reducing PR should lead to a reduction in other anti-masks attitudes. Researchers in the field of mass communication have suggested several strategies for improving the persuasiveness of messages in situations in which PR might occur. Indirect or subtle types of messaging—known as “nudges” in behavioral economics (62)—could be used. A nudge is something that alters a person’s behavior in a predictable way without forbidding any options or significantly changing their economic incentives. Mask-related nudges could consist of handing out free masks at the entrances to store or displaying brightly colored posters of happy cartoon characters wearing masks. It is unclear whether such nudges are effective when PR plays a role in mask refusal.

Other forms of messaging have been proposed as ways of specifically dealing with PR. A review of the research literature on mass communication (48) concluded that several messaging strategies could circumvent PR. These strategies could be adapted for mask-related PR, as follows:

a. Add postscripts to messages that emphasize freedom of choice (e.g., “Please do your part in managing the pandemic by wearing a mask. The choice is yours.”). The postscript is intended to ameliorate PR.
b. Narratives that highlight personal choice (e.g., personal stories about why people who were initially reluctant to wear masks eventually chose to wear masks).
c. Messaging that emphasizes how an individual’s choices impact others (e.g., “Choosing to wear a mask shows that you care about your community”).
d. Messages that on the surface address one audience but are really targeted at a different audience who may be listening (e.g., “Thank you for choosing to wear masks. Wearing a mask might seem to be a small thing to it, but it is vitally important.”).
e. Messages that forewarn receivers about the potential of them experiencing reactance (e.g., “Some people think they’re giving up freedom by wearing a mask. But that’s not true. Wearing a mask is a way of freeing ourselves from the pandemic”).
f. Using reactance to strengthen the message (e.g., “You have a right to wear a mask to stay safe. Don’t let anyone take away your right”). This message could help mask-refusers to realize that wearing a mask is a right that they can choose to adopt.

The efficacy of these messaging strategies for improving mask adherence remains to be investigated. No single message is effective under all circumstances, and pilot testing is required before messages are implemented in mass communication programs in order that they be fine-tuned and to avert any unintended adverse effects of messaging.

The present study has various strengths and limitations. In terms of strengths, the sample was large and the present study was, to our knowledge, the first to use network analysis to understand the interrelationships among anti-mask attitudes and their relationships to other variables that have been shown to influence attitudes and behavior in the context of pandemics. A further strength of the present study was that, compared to previous surveys, the present study examined a larger number of different types of anti-mask attitudes as they related to mask-wearing and other variables. A limitation is that not all possible anti-mask attitudes were assessed. Future research is needed to investigate anti-masks attitudes that were not investigated in the present study. For example, the present study did not assess the belief that mask-wearing is a sign of weakness, which was an attitude voiced in an Ohio anti-mask rally (63). The replicability of the findings across different countries and cultures also remains to be investigated in future research. A further limitation is that network analysis, as a statistical modeling method, is insufficient for determining the causal nature of the relationships between nodes. Nevertheless, the present findings provide a strong rationale for conducting future experimental studies on the causal status of mask-related PR and beliefs that masks are ineffective. The present study also underscores the importance of conducting future research to investigate the efficacy of messaging strategies that take PR into consideration as a means of addressing the problem of mask non-adherence.

## Data Availability

Data are available on request.

## Acknowledgements

The authors thank Michelle M. Paluszek, Caeleigh A. Landry, and Geoffrey S. Rachor for their assistance in completing this study.

